# Associations between intimate partner violence and mental health in German men and women: a cross-sectional analysis of the German Health Interview and Examination Survey for Adults (DEGS1)

**DOI:** 10.1101/2021.12.20.21268089

**Authors:** Lena Graßkemper, Diogo Costa

## Abstract

This work explores the cross-sectional associations between Intimate Partner Violence (IPV) and anxiety, depressive symptoms, stress symptoms, and health-related quality of life (HRQoL), in a representative sample of German adult men (n=2,789) and women (n=3,149), and considers their involvement as victims or perpetrators of physical and psychological IPV. In this sample, physical IPV victimization was associated with anxiety and stress among men. Psychological IPV victimization was associated with depression among men, and with stress among both sexes. Physical IPV perpetration was significantly associated only with women’s depressive and stress symptoms. Psychological IPV perpetration was associated with stress for both men and women. The mental component of HRQoL was significantly lower for men and women involved in any type of IPV. These results support the need to consider the mental health consequences of IPV involvement for both men and women.

## Introduction

Intimate partner violence (IPV) is one of the most frequently experienced forms of violence and is a significant and widespread problem around the world (Garcia-Moreno, Jansen, Ellsberg, Heise, & Watts, 2006). Internationally, IPV is recognized as one of the “most prevalent human rights violations in the world” (United Nations Population Fund [UNFPA], 2020, para. 1) and it has been considered as a global public health problem of epidemic proportions (Campbell, 2002; Ellsberg, Jansen, Heise, Watts, & Garcia-Moreno, 2008; World Health Organization [WHO], 2013). The World Health Organization (WHO) defines behaviors in an intimate relationship as IPV as soon as they involve physical, sexual or psychological harm (Butchart, Garcia-Moreno, & Mikton, 2010).

Approximately one in three women all over the world have been a victim of IPV. Globally, the lifetime prevalence of physical and/or sexual IPV violence since the age of 15 is 26% among ever-married or partnered women and is about 10% when considering the 12-month prevalence (World Health Organization, 2021). The prevalence in Europe was found to be 33% (European Union Agency for Fundamental Rights [FRA], 2014). As a recent review observed, IPV is more prevalent during a woman’s lifetime than other conditions such as diabetes or breast cancer and it is more widespread than common mental health conditions (Miller & McCaw, 2019). Research mostly focused on women as victims of IPV (Frieze, Newhill, & Fusco, 2020; Garcia-Moreno et al., 2006). Nevertheless, some studies about men victimization are available. One review, including 91 studies, observed that one in five men had been a victim of IPV (Desmarais, Reeves, Nicholls, Telford, & Fiebert, 2012). Data from Germany showed that every second or third woman had ever experienced physical violence, while every fifth to seventh woman had suffered physical or sexual abuse at the hands of her partner (Bundesministerium für Familie, Senioren, Frauen und Jugend [BMFSFJ], 2004b). Furthermore, one in four of 200 German men surveyed in a qualitative study experienced at least one act of physical violence from their intimate partner and psychological violence was the most frequently reported type (BMFSFJ, 2004a). However, the real extent of IPV is hindered by lack of data in many countries, as IPV is occurring in the private setting and only a small percentage of victims seek help from service providers or report cases to official authorities (Langhinrichsen-Rohling, 2005; WHO, 2014).

IPV can lead to deleterious consequences of the individuals involved and is a major contributor to various health problems (Ellsberg et al., 2008), including several mental health outcomes, for both men and women (Lagdon, Armour, & Stringer, 2014). A link between anxiety and IPV perpetration has been previously documented (Barbaro, Boutwell, & Shackelford, 2019; C. Spencer et al., 2019; C. M. Spencer, Keilholtz, & Stith, 2021), and depression is one of the most studied consequences of IPV (Frieze et al., 2020; Warshaw et al., 2009) found among women snd among men (Devries et al., 2013; Pengpid & Peltzer, 2020; Randle & Graham, 2011; Romito, Beltramini, & Escribà-Agüir, 2013). Studies that examined the link between IPV perpetration and depression are less frequent, but also showed relevant associations (Fulu et al., 2013; Ulloa & Hammett, 2016).

Women and men with a history of IPV have been found to present lower scores of health-related quality of life (HRQoL) than women not experiencing IPV (Beeble, Bybee, Sullivan, & Adams, 2009; Bonomi et al., 2006; Dillon, Hussain, Loxton, & Rahman, 2013; Reid et al., 2008) and the association may depend on the type of involvement, as victim or perpetrator (Costa, Hatzidimitriadou, et al., 2015). Few studies have captured stress as a consequence of IPV experiences, although there is evidence suggesting that IPV is associated with psychological stress (Yim & Kofman, 2019) and with posttraumatic stress disorder (Pico-Alfonso et al., 2006; Seedat, Stein, & Forde, 2005).

The physical and mental health consequences have often been reported in the scientific literature but focus primarily on victims (Lagdon et al., 2014; Pengpid & Peltzer, 2020; Próspero, 2007; Williams, Murphy, Dore, Evans, & Zonderman, 2017). Fewer studies have focused on the consequences for the perpetrator’s health, including mental health, or the influence of psychological IPV (Heise et al., 2019). Moreover, most of the studies assessed the consequences of IPV among women and considered mainly clinical or selected victim samples (Afe, Emedoh, Ogunsemi, & Adegbohun, 2016; Bonomi et al., 2006).

The consequences of involvement in IPV as a perpetrator, may also be different for men and women, suggesting different etiologies for mental health problems worth of future explorations. For example, women may show depressive symptoms that result from feelings of guilt, shame or regret for being a perpetrator (Graham, Bernards, Flynn, Tremblay, & Wells, 2012), while this might not be observed among men.

This study aims to assess the associations between physical and psychological IPV and the mental health of German adult men and women, considering their involvement as victims or perpetrators, while controlling for known influential factors. For this purpose, we resort to the German Health Interview and Examination Survey for Adults (DEGS1) study population (2008-2011) and to four important mental health outcomes: anxiety disorder, depression symptoms, stress symptoms, and health-related quality of life. Although related, the study of these separate outcomes can enlighten about the differential impact that involvement in each IPV type can have on exposed individuals and inform targeted interventions (e.g., towards prevention of anxiety or mood disorders, stress, or the physical dimension of quality of life).

## Methods

### Sample and Study Participants

The present cross-sectional study was based on data obtained from the first wave (2008-2011) of the German Health Interview and Examination Survey for Adults (DEGS1), conducted by the Robert Koch-Institute (RKI) (Robert Koch-Institute Department of Epidemiology and Health Monitoring, 2015). The concept and design, sampling and response rates of the DEGS1 study are described in detail elsewhere (Gößwald, Lange, Kamtsiuris, & Kurth, 2012; Kamtsiuris et al., 2013; Scheidt-Nave et al., 2012). Briefly, the survey included self-completion written questionnaires, physical examinations and tests of physical and cognitive functions, using validated methods and standardized tools for the German population. The target population comprised resident adults aged 18–79 years. The study was designed to allow both cross-sectional and longitudinal types of analyses. To do so, a random sample from local population registries was drawn to complete the participants of the, previously established, German National Health Interview and Examination Survey 1998 (GNHIES98), who re-participated. A total of 8,152 adults (3,869 men; 4,283 women) aged 18-79 years with a permanent residence in Germany participated, including 4,193 first-time participants (response rate 42%) and 3,959 revisiting participants of the GNHIES98 (for response rate 62%) (Scheidt-Nave et al., 2012). Since experiences of physical and psychological violence were only assessed in the age range from 18-64 years (Kamtsiuris et al., 2013), participants with the age of 65 years and older were excluded from the current analyses. Accordingly, this paper includes 5,938 participants, of whom 2,789 (47.97%) were men and 3,149 (53.03%) were women.

## Measures

### Intimate partner violence - IPV

In the DEGS1 study a specific screening instrument was developed and used for the assessment of violence and the details of development can be found elsewhere (Lange, Starker, Lippe, & Hölling, 2016; Schlack, Rüdel, Karger, & Hölling, 2013). The questionnaire assessed IPV experiences together with family violence, violence at the workplace, violence experiences with other known persons and with unknown persons. All the questions assessing an involvement in violence referred to the previous 12 months. We analyzed the questions about physical and psychological violence and considered answers concerning victimization and perpetration experiences.

To assess the involvement in physical violence victimization, participants were asked if they had experienced somebody physically attacking him/her. In order to define a physically attack, examples were given: hitting, slapping, pulling hair, kicking or threatening with a weapon or an object. The response options were “yes” and “no”. If participants answered “yes”, they were asked about the perpetrator of the attack (“partner”, “family/relatives”, “colleagues/bosses”, “known person”, “unknown person”). Multiple answers were permitted. For the assessment of perpetration experiences, participants were asked if they had ever attacked someone physically. Likewise, the same definition was given, and possible outcomes were “yes” and “no”. If participants answered “yes”, they were asked to elaborate about the victim of their violent act. Possible choices were the same than given before and answers could be multiple. In this study, IPV was considered when victims/perpetrators identified involvement in violence with the partner.

The questions about psychological violence also comprised a definition of psychological violence: “[…] has any other person been derogatory towards you (for example, with regard to your appearance, the way you dress, think, act or work or any possible disabilities)? Or have you been insulted, badmouthed, threatened, bullied or put under pressure?”. Similarly, possible outcomes were binary (“yes”/”no”), and if participants answered “yes”, they were asked further about the perpetrator of the attack (“partner”, “family/relatives”, “colleagues/bosses”, “known person”, “unknown person”; multiple answers were permitted). Regarding perpetration of psychological violence, participants were asked if they have been psychologically offensive toward any other person, and if so, towards whom. This question contained the same definition of psychological violence as given above, and possible conflict persons of the psychological attacks were also the same groups as indicated before. Multiple answers were permitted. IPV was considered when victims and perpetrators declared involvement in violence with the partner.

### Mental Health

In this analysis the outcomes anxiety disorder, depressive symptoms, stress and quality of life were used to describe the mental health of the participants. For the assessment of anxiety disorders, study participants were asked if they suffer or ever suffered from an anxiety disorder which was diagnosed by a physician. Possible outcomes were binary (“yes”/”no”). When a diagnosis was present, further questions followed about the age in which a diagnose was first made and if study participants had an anxiety disorder within the last 12 months (12-month prevalence). For further analyses, the 12-month prevalence was used in this paper.

For the assessment of depressive symptoms, the depression module of the PHQ-D were used (composed of nine items). The PHQ-D items referred to feelings, situations and impairments in the previous two weeks and could be answered with a 4-point Likert Scale, which asked how often the respective situation was experienced (0 never; 1 on single days; 2 on more than half of the days; 4 almost every day) (Scheidt-Nave et al., 2012). The sum of the scores across the nine items could range from 0 to 27. The outcome “depression” was categorized, with a distinction between “no current depressive symptoms” (0-9 points) and “current depressive symptoms” (10-27 points). This distinction was chosen according to the literature (Busch, Maske, Ryl, Schlack, & Hapke, 2013; Löwe, Spitzer, Zipfel, & Herzog, 2002).

The short version of the Trier Inventory for Chronic Stress, named Screening Scale of Chronic Stress (SSCS) was used to assess participants’ chronic stress (Scheidt-Nave et al., 2012). It consists of 12 items and measures the frequency of subjectively experienced stress in the five following stress domains in the last three months: chronic preoccupation, work-related and social overload, excessive demands, and lack of social recognition. The frequency of stress in each of the five stress areas was asked with a 5-point Likert Scale (0 never; 1 rare; 2 sometimes; 3 often; 4 very often). The 12 items were summed to a scale sum score, ranging from 0 and 48 points (Schulz, Schlotz, & Becker, 2004). The outcome stress was categorized based on the average. Accordingly, participants were divided into two groups: “below-average to average” with 0-11 points or “above-average” with 12-48 points. The choice of the cut-off point was determined based on the literature (Hapke et al., 2013).

To assess the HRQoL, the German Version of the Short Form-36 Health Survey Version 2 (SF-36) was used. The SF-36 consists of 36 items that yield in 8 dimensions of subjective health and refers to the previous four weeks. The dimensions are physical functioning, physical role functioning, bodily pain, general health, vitality, social functioning, emotional role functioning and mental health. Participants had to answer with 3- or 5-points Likert Scales. The resulting eight subscales were categorized into two basic dimensions: the physical and the mental component summary score for HRQoL (Morfeld, Kirchberger, & Bullinger, 2011). The categorization of the 36 items into two basic dimensions is described in detail elsewhere (Ellert & Kurth, 2013; Ware et al., 2007). In brief, both scales were normalized with a mean=50 and a standard deviation=10. The higher the scores, the better was the expected HRQoL.

### Covariates

Age, marital status, general health status, socio-economic status (SES) and childhood abuse were included as covariates in this study. For descriptive analysis, age was divided into five age-groups: 18-24; 25-34; 35-44; 45-54; 55-64 years. The marital status was coded as: married (same household as partner), married (different household as partner), single, divorced and widowed. The general health status was assessed with five categories and categorized into two: “very good/good” and “moderate/poor/very poor”

For the assessment of the SES an index was used, including information on formal education and vocational training, occupational status and net household income. Each dimension could be scored from 1 to 7, with the SES index corresponding to the sum of the dimensions, hence taking values between 3 and 21. Low values represented a low SES. The categories used and associated point values are described in detail elsewhere (Lampert, Kroll, Müters, & Stolzenberg, 2013). In the current analysis the quintiles of the distribution of the SES index were used.

As childhood abuse by the parents has been identified as a risk factor for the involvement in IPV (Godbout et al., 2019; Lang, Stein, Kennedy, & Foy, 2004; Stermac, Reist, Addison, & Millar, 2002) and is linked with mental health outcomes (Bandelow et al., 2004), participants were asked about their experiences of physical assault by their parents/guardians before their 16^th^ birthday. Answer options were “frequently”, “occasionally”, “rarely” and “never”.

### Ethical considerations

The DEGS1 study was consented with the Federal and State Commissioners for Data Protection and approved by the Medical Ethics Review Committee of the Charité in Berlin in September 2008 (Scheidt-Nave et al., 2012).

The survey was conducted in health examination centers and questions addressing IPV were self-administered. The study staff was continuously trained and supervised with regard to the survey content. Local crisis lines and emergency addresses were available for each examination team in case of re-traumatization potentially caused by the assessment of violence (Schlack et al., 2013).

### Statistical analysis

Frequencies and means were computed to describe participants characteristics. A weighting factor was used in analysis to correct sample deviations from the population structure for age, sex, region, nationality, type of municipality and education (Kamtsiuris et al., 2013). Complex Samples commands were used in SPSS v26 to accommodate weighting and clustering of participants within sample points in the calculation of p-values and confidence intervals.

To test differences in the categorical outcomes (anxiety disorders, depressive symptoms and stress level), Chi-square and Fisher Exact tests were performed. T-tests were used to compare means of the SF-36 summary components according to IPV involvement. All comparisons, in which participants involved in IPV were compared to participants not involved in IPV, were differentiated by type of involvement (victim, perpetrator) and type of violence (physical, psychological), and stratified by sex. Participants who declared involvement in one specific type of IPV (e.g. physical IPV perpetration) were compared to participants who declared not being involved in this specific type of IPV (e.g. no physical IPV perpetration).

Logistic regression (categorical outcomes) and linear regression models (SF-36 summary components) were fitted to examine the associations between IPV and the mental health outcomes. The models were stratified according to the type of violence (physical, psychological), involvement in IPV (victim, perpetrator) and by sex. In case of logistic regressions, models were adjusted for age, marital status, general health status, socio-economic status (SES) and childhood abuse, and the magnitude of associations expressed as odds ratios and respective 95% confidence intervals (CI). For the linear regression, β coefficients and standard errors and 95% CI for the associations between IPV involvement and HRQoL were computed, in models adjusted for age, marital status, socio-economic status (SES) and childhood abuse. A p-value of <0.05 was considered as significant.

## Results

### Descriptive Statistics

Most of the participants were 45-54 years old (25.9%) and married (same household 58.1%). Over half of the study participants had an intermediate SES (2^nd^-4^th^ quintile: 60.7%) and the majority of the participants described their subjective general health status as very good or good (79.4%). Overall frequency of anxiety was 3.1%. 8.5% of the study population had increased depressive symptoms and 48.9% an increased stress level. The mean score (95% confidence intervals) in the physical and mental components of the SF-36 were 52.66 (52.38-52.93) and 48.89 (48.54-49.24) respectively.

### IPV Involvement and Mental Health

IPV victimization rates were higher for women than for men (see Tab. 2a & 2b). A total of 0.9% of the men and 1.2% of the women had reported physical IPV victimization within the last 12 months, whereas 2.8% of the men and 5.2% of the women reported psychological IPV victimization within the last 12 months. However, declared IPV perpetration rates were higher among women than men (see Tab. 2a & 2b). 0.3% of the men stated that they perpetrated physical IPV against their partner in the last 12 months, while 1.3% of the women reported physical IPV perpetration. Also, more women (3.6%) declared that they perpetrated psychological IPV against their partner compared to men (2.6%).

Men involved in IPV had a worse mental health than those declaring no IPV experiences (Tab 2a). Men victims and perpetrators of physical and psychological IPV had significantly more often an anxiety disorder diagnosis than participants not involved in IPV (p<0.05). Also, significantly more men involved in IPV with current depressive symptoms, an above average stress level and a significantly lower mental component summary score were observed compared to men who reported not being involved in IPV. Women involved in IPV were also observed to have worse mental health outcomes than women not involved in IPV (Tab. 2b). Significantly more women victims of physical IPV suffered from anxiety disorders than women without IPV experiences (10.2%, vs. 4.0%, p<0.05). Furthermore, regardless of type of involvement or violence, women with IPV experiences had more often depressive symptoms and an above-average stress level than women without IPV experiences. The mental QoL component summary score was significantly lower among women involved in IPV for all types of violence and involvement compared to women without involvement, with the biggest difference for victims of physical violence (38.92; SE=2.75 vs. 47.78; SE=0.25; p<0.001). Significant differences in the physical component summary score were only found in the group of women declaring perpetration of physical IPV compared to women declaring no perpetration of physical IPV (p<0.05).

### Associations between IPV and Mental Health Outcomes

#### Anxiety disorders

Men participants who reported physical IPV as victims, were significantly more likely to report an anxiety disorder compared to men not declaring physical IPV victimization after adjustment for age group, marital status, SES, general health and childhood abuse experiences (aOR=12.23; 95%-CI=2.29-65.40), Tab 3. For women, no significant associations were found between IPV involvement and anxiety.

#### Depressive symptoms

Men victims of psychological IPV presented increased odds of having current depressive symptoms compared to men not reporting IPV psychological victimization (aOR=3.85; 95%-CI=1.87-7.92). Women who declared perpetration of physical IPV also presented higher odds of depressive symptoms (aOR=3.72; 95%-CI=1.34-10.30) compared to women that did not report physical IPV perpetration.

#### Stress

Results suggested strong associations between the involvement in IPV and an above-average stress level for both men and women (Tab 3). Men victims of physical IPV presented a significantly higher chance of having an above average stress level compared to men participants without physical IPV victimization experiences (aOR=6.21; 95%-CI=1.42-27.28). Both men and women victims of psychological IPV had higher odds of an increased stress level in comparison to participants without psychological IPV experiences (men: aOR=3.91; 95%-CI=1.98-7.73; women: aOR=2.04; 95%-CI=1.23-3.38). Men and women participants declaring psychological IPV perpetration were also found to have an above average stress level compared to those not declaring perpetration (men: aOR=5.49; 95%-CI=2.57-11.73; women: aOR=2.76; 95%-CI=1.33-5.70).

#### Health-related Quality of life

Regarding the physical summary component of the SF-36, a significant positive association was observed for men declaring psychological IPV perpetration (β=2.536; SE=3.046) and for women declaring physical IPV perpetration (β=2.434; SE=1.181), suggesting higher scores (better HRQoL in this dimension) for these groups (Tab 4).

Statistically significant negative associations with the mental summary component of the SF-36 were observed among men and women who declared physical (β=-4.667; SE=1.329, and β=-6.905; SE=2.599, respectively) and psychological IPV victimization (β=-6.772; SE=1.270, and β=-6.426; SE=1.181, respectively). The same was noted in this component for women who reported physical IPV perpetration (β=-7.231; SE=2.292), and for men and women who reported psychological IPV perpetration (β=-5.379; SE=1.431 in men, and β=-7.769; SE=1.623 in women).

## Discussion

This study aimed to examine the associations between IPV involvement and selected mental health outcomes (anxiety, depressive symptoms, stress and HRQoL), among German adult men and women. The results showed that women, compared to men, declared more frequently to be involved in physical and psychological IPV as victims and as perpetrators, in the previous 12 months. Furthermore, the results suggest that participants involved in IPV had significantly poorer mental health than participants not reporting IPV involvement, independently of several potential confounders (age, marital status, SES, general health status and experiences of childhood abuse).

## Prevalence

The prevalence observed in our study for IPV victimization was lower when compared to international studies, such as the European DOVE study (suggesting a prevalence of 13.5% for women and 12.3% for men victims of physical IPV in Stuttgart, Germany) and the FRA-study (suggesting a prevalence of 3% for women victims of physical IPV in Germany) (Costa, Soares, et al., 2015; FRA, 2014). Higher victimization rates for women in Germany were also found in the national study on violence against women, for physical IPV victimization, 8% for the age group 16-to 49-year old women, and 3% for the 50-to 65-year old group of women (Stöckl & Penhale, 2015). The varying victimization rates could be explained by the different methods and survey instruments used, since one overall approved and acknowledged method for the assessment of IPV does not exist, thus hindering comparisons. The same can be said regarding the perpetration measurement, although few evidence exists, also pointing to higher rates (Costa, Hatzidimitriadou, et al., 2015; Fulu, Jewkes, Roselli, & Garcia-Moreno, 2013).

The similarities in the frequency of IPV victimization and perpetration found for men and women, can be a result of the evolving gender equality of western societies as postulated by theories of social roles (Archer, 2009). Self-defense is also considered as an explanatory reason for higher perpetration rates found among women. It is argued that women primarily use violence against a male partner in self-defense (Kimmel, 2002) and as resistance to their partner’s controlling behavior (Johnson, 2006). Because of the broad variety of explanatory approaches, researchers point out that women perpetration towards their partner cannot be understood using a unitary explanation (Graham-Kevan & Archer, 2005). The DEGS1 study did not assess contextual relevant factors (e.g., controlling behaviors), which could have helped to explain the gender differences in perpetration. This should be the focus of further research efforts in the German society.

## Mental Health Outcomes

The results of this paper suggested that IPV involvement is associated with poorer mental health outcomes for both men and women. Considering men, mixed findings have been previously documented. Some studies found significant correlations between IPV experiences and anxiety disorders in men, in line with our results (Lagdon et al., 2014; Shorey et al., 2011). While others did not found significant differences (Próspero, 2007; Romito et al., 2013). Regarding women, we could not find significant associations between IPV involvement and anxiety disorders in, although this has been found in previous studies (Afifi et al., 2009; Dillon et al., 2013; Romito et al., 2013). The outcome “anxiety disorder” analyzed in this study referred to the last 12 months and was considered only if it had ever been medically diagnosed. Thus, women could have experienced anxiety symptoms because of their involvement in IPV, but this was measured or diagnosed in our sample, which could have led to an underestimation of anxiety symptoms prevalence and their association with IPV. An anxiety disorder may also develop later as a long-term consequence of IPV, thus the 12-month prevalence assessed in this study would not capture an anxiety disorder as a possible long-term consequence of IPV.

Women involved in IPV, showed higher odds of having depressive symptoms compared to women participants who declared not being involved in IPV. Several studies and reviews are in line with our results and also observed a higher likelihood of depressive symptoms among women with a history of IPV (Dillon et al., 2013; Lagdon et al., 2014; Wood, Voth Schrag, & Busch-Armendariz, 2020). In line with other studies (Barker, Stewart, & Vigod, 2019; Randle & Graham, 2011; Reid et al., 2008), we found a significant relationship between victimization and depressive symptoms among men. The association between IPV perpetration and elevated depressive symptoms was also observed among women reporting physical IPV perpetration, but not among men. It has been suggested that perpetration of physical violence acts may lead women to feelings of shame and guilt for their behavior, which can then translate into depressive symptoms (Graham et al., 2012), while the same is not observed in men. Additionally, specific gender roles could complicate the expression of depressed mood and the seeking of help for depression (Möller-Leimkühler, 2008), which could lead to an underrepresentation of the real extent in this data, particularly among men.

A comparison of the stress scores with other studies turned out to be difficult because we could not identify previous studies using the SSCS questionnaire and examining it in relation to IPV. Furthermore, the stress level was only related to the average stress level of the underlying study population. Consequently, it hinders comparisons with other study populations. Nonetheless, the results found are in line with a recent systematic review showing strong evidence that IPV is associated with psychological stress (Yim & Kofman, 2019). Since stress has been observed to be also a significant risk factor for other mental or physical illnesses (Russ et al., 2012), the results of this study highlight the increased risk of participants involved in IPV for further negative health consequences.

In line with other international studies, we did not found a clear connection between a decreased physical component summary score and IPV involvement in men and women (Bonomi et al., 2006; Costa, Hatzidimitriadou, et al., 2015; Reid et al., 2008). In contrast, strong associations were observed between IPV and a lower mental component summary score. Accordingly, the results suggested that IPV primarily impacts the mental QoL rather than the physical QoL. It can be assumed that the physical consequences affect the victim and perpetrator rather short-term and do not entail long-term limitations. This explanation is supported by available data from the FRA-study: more than half of the victims of IPV did not suffer any injuries, while the other half mostly suffered only small injuries (FRA, 2014). However, other studies also found a significant associations between the mental health dimension of QoL and IPV for both men and women (Bonomi et al., 2006; Costa, Hatzidimitriadou, et al., 2015; Dillon et al., 2013; Reid et al., 2008), which is in agreement with this paper.

## Strengths and limitations

The strengths of this study included the analysis of a large population-based national representative sample of the adult population. This research contributes to existing literature by suggesting a mental health impact of psychological and physical IPV involvement among women and men. It was possible to analyze the health effects of IPV while simultaneously controlling for potential confounders. Furthermore, the DEGS1 study made use of highly standardized and validated questionnaires in order to measure the mental health outcomes. The use of these tools allows for comparisons with other findings. For Germany, this paper represents an important addition to the current state of research on violence and mental health. The DEGS1 surveyed physical and psychological violence events for men and for women as perpetrators and victims for the first time on a nationally representative level, making it possible to analysis gender-specific impacts to different mental health outcomes.

A major limitation lies in the violence questionnaire. Experiences of sexual IPV and controlling behaviors were not surveyed and no differentiation was made between violence committed (on a one-time basis) in the heat of the moment and systematic violence committed on a regular basis (chronicity of acts). Furthermore, the use of a non-standardized instrument to assess IPV, that does not provide information about violence severity or the chronicity of acts, limits comparability with other international studies. Nevertheless, since the DEGS1 screening instrument was tested successfully (Schlack et al., 2013), and its contents are comparable to items composing the Revised Conflict Tactics Scale (CTS2), which is one of the most commonly used IPV assessment tools (Jones, Browne, & Chou, 2017; Straus & Douglas, 2004), we believe these results retain their validity.

The main DEGS1 study was conducted using self-completion written questionnaires. However, different survey methods can influence the disclosure of experiences of IPV, which can lead to an underestimation of the extent of the problem (Mirrlees-Black, 1999). The availability and willingness of the individual to recognize their involvement in IPV, as well as the modalities of asking questions, are further influencing factors on data availability (Fraga, 2016). Because of the sensitive and multifaceted nature of IPV, low disclosure rates cannot be discarded. Moreover, since some of the questions referred to the previous 12 months, the risk of a recall bias also emerges. The results are based on retrospective recall which can lead to an over- or underestimation of the events. Also, a reporting bias due to social desirability responses cannot be ignored.

A gender bias may have impacted the self-reports of violent behavior. Within male and female role models, there are different interpretations and perceptions of physical confrontations. If men perpetrate acts of violence, these acts are more likely to be accepted and tolerated, especially from persons of the same sex, as a constituent part of the male role model (Meuser, 2002). Women’s perpetration, on the other hand, is in contradiction with cultural expectations and gender roles. Some acts might therefore be more quickly interpreted as violent acts within the female gender role, while acts of male perpetration are accepted as more normal within the male gender role (Gilbert, 2002).

Even though the data of the DEGS1 study was collected before the COVID-19 pandemic, recent findings showed that the topic needs to gain more attention. Countries all over the world have implemented various restrictions for the containment of the SARS-CoV-2 virus, which involves strict stay-at-home orders. In one aspect, these measures help to slow down the spread of COVID-19 but on the other hand may lead to an increase in the occurrence of IPV (Jetelina, Knell, & Molsberry, 2021; Matoori et al., 2020). Researchers found that all EU Member States implemented changes or established new support and protection services for victims of IPV and their children in response to COVID-19. Still, only a few EU Member States implemented comprehensive action plans specifically addressing the issue of IPV in the context of the COVID-19 pandemic (European Institute for Gender Equality, 2021). It becomes apparent, that, while stopping the spread of the COVID-19, the epidemic of intimate partner violence cannot be ignored.

## Conclusions

The experience of IPV victimization and perpetration is strongly associated with poor mental health outcomes. Regardless of sex, involvement as victim or perpetrator, and type of violence (physical or psychological), people involved in IPV had higher odds of suffering from an anxiety disorder, depression, increased stress levels or decreased HRQoL compared to people who were not involved in IPV. It becomes crucial to provide gender-sensitive services to each affected individual with the aim to mitigate the negative consequences of IPV.

## Data Availability

The DEGS1 dataset (German Health Interview and Examination Survey for Adults) is accessible on application to interested researchers as anonymized data for secondary analysis.

## Grant numbers and/or funding information

The DEGS1 study is done by the RKI as part of the Federal Health Monitoring in Germany on behalf of the Federal Ministry of Health. The funders had no role in study design, data collection and analysis, decision to publish, or preparation of the manuscript.

## Tables

**Table 1:** Social and demographic characteristics of total study population (n=5,938)

| Characteristics | n | %<br>Weighted | 95%<br>Confidence<br>Intervals |
| --- | --- | --- | --- |
| **Sex** |  |  |  |
| male | 2789 | 50.6 | 48.9-52.2 |
| female | 3149 | 49.4 | 47.8-51.1 |
| **Age** |  |  |  |
| 18-24 | 630 | 13.1 | 12.2-14.0 |
| 25-34 | 917 | 19.0 | 17.7-20.4 |
| 35-44 | 1255 | 22.5 | 21.2-23.9 |
| 45-54 | 1684 | 25.9 | 24.6-27.2 |
| 55-64 | 1452 | 19.5 | 18.5-20.7 |
| **Marital status** |  |  |  |
| married, same household | 3567 | 58.1 | 56.2-60.0 |
| married, diff. households | 105 | 2.1 | 1.6-2.6 |
| single | 1619 | 31.5 | 29.9-33.1 |
| divorced | 407 | 6.1 | 5.5-6.9 |
| widowed | 143 | 2.2 | 1.8-2.7 |
| **Socioeconomic status** |  |  |  |
| 1<sup>st</sup> quintile | 847 | 18.0 | 16.6-19.6 |
| 2<sup>nd</sup> quintile | 1026 | 19.2 | 17.9-20.6 |
| 3<sup>rd</sup> quintile | 1182 | 20.0 | 18.7-21.3 |
| 4<sup>th</sup> quintile | 1313 | 21.5 | 20.2-22.7 |
| 5<sup>th</sup> quintile | 1506 | 21.4 | 19.7-23.1 |
| **General health** |  |  |  |
| very good/good | 4658 | 79.4 | 78.0-80.6 |
| moderate/(very) poor | 1219 | 20.6 | 19.4-22.0 |
| **Childhood abuse** |  |  |  |
| never | 3579 | 62.3 | 60.7-64.0 |
| rarely | 1386 | 24.2 | 22.7-25.7 |
| occasionally | 548 | 9.7 | 8.8-10.7 |
| frequently | 195 | 3.8 | 3.1-4.6 |
| **Mental health** |  |  |  |
| Anxiety yes | 160 | 3.1 | 2.5-3.8 |
| no | 5712 | 96.9 | 96.2-97.5 |
| Depression<sup>1</sup> yes | 452 | 8.5 | 7.6-9.6 |
| no | 5219 | 91.5 | 90.4-92.4 |
| Stress<sup>2</sup> yes | 2813 | 48.9 | 47.2-50.6 |
| no | 2988 | 51.1 | 49.4-52.8 |
|  | **n** | **mean** | **95%-CI** |
| HRQoL Physical |  | 52.66 | 52.38-52.93 |
| Mental |  | 48.89 | 48.54-49.24 |

<sup>1</sup>PHQ-9 score depressive symptoms: yes > 9; no ≤ 9
<sup>2</sup>SSCS-score: stress level: yes > 11, no ≤ 11
HRQoL: Health-related Quality of Life, summary components from the SF-36;

**Table 2a:** Mental health outcomes of men involved in intimate partner violence (IPV) as victims and perpetrators compared to men not involved in IPV, according to type of violence (physical, psychological)

| Outcome | Victim |  |  |  | Perpetrator |  |  |  |
| --- | --- | --- | --- | --- | --- | --- | --- | --- |
|  | Physical |  | Psychological |  | Physical |  | Psychological |  |
|  | IPV | No IPV | IPV | No IPV | IPV | No IPV | IPV | No IPV |
|  | n (%) | n (%) | n (%) | n (%) | n (%) | n (%) | n (%) | n (%) |
| Total | 18 (0.9) | 2710 (99.1) | 77 (2.8) | 2658 (97.2) | 10 (0.3) | 2726 (99.7) | 63 (2.6) | 2672 (97.4) |
| Anxiety yes | 3 (21.9) | 40 (1.9) | 4 (5.1) | 38 (1.9) | 2 (27.6) | 41 (1.9) | 5 (5.5) | 38 (1.9) |
| no | 15 (78.1) | 2650 (98.1) | 72 (94.9) | 2601 (98.1) | 8 (72.4) | 2664 (98.1) | 57 (94.5) | 2614 (98.1) |
| p* |  | <0.05 |  | <0.05 |  | <0.05 |  | <0.05 |
| Depression<sup>1</sup> yes | 5 (36.2) | 155 (6.0) | 16 (23.3) | 145 (5.8) | 2 (22.4) | 159 (6.2) | 7 (9.2) | 154 (6.3) |
| no | 13 (63.8) | 2481 (94.0) | 60 (76.7) | 2438 (94.2) | 7 (77.6) | 2492 (93.8) | 56 (90.8) | 2442 (93.7) |
| p* |  | <0.05 |  | <0.05 |  | 0.099 |  | 0.102 |
| Stress<sup>2</sup> yes | 14 (86.3) | 1148 (43.7) | 59 (78.1) | 1102 (42.9) | 8 (84.2) | 1155 (43.8) | 49 (81.1) | 1115 (42.9) |
| no | 4 (13.7) | 1540 (56.3) | 18 (21.9) | 1534 (57.1) | 2 (15.8) | 1549 (56.2) | 14 (18.9) | 1536 (57.1) |
| p* |  | <0.05 |  | <0.001 |  | <0.05 |  | <0.001 |
|  | mean (SE) | mean (SE) | mean (SE) | mean (SE) | mean (SE) | mean (SE) | mean (SE) | mean (SE) |
| HRQoL Physical | 52.35 (2.52) | 53.09 (0.21) | 52.03 (1.31) | 53.12 (0.21) | 54.16 (3.64) | 53.10 (0.21) | 55.82 (0.91) | 53.02 (0.22) |
| p** |  | 0.548 |  | 0.278 |  | 0.646 |  | 0.096 |
| HRQoL Mental | 42.44 (1.77) | 50.13 (0.24) | 41.87 (1.37) | 50.32 (0.23) | 39.44 (3.76) | 50.11 (0.24) | 43.31 (1.45) | 50.25 (0.24) |
| p** |  | <0.05 |  | <0.001 |  | <0.05 |  | <0.001 |

n=unweighted count; % or mean= weighted estimate;
\*p-values from Chi-square or Fisher Exact test;
\*\*p-values from T-test;
<sup>1</sup>PHQ-9 score depressive symptoms: yes > 9; no ≤ 9;
<sup>2</sup>SSCS-score stress level: yes > 11; no ≤ 11;
HRQoL: Health-related Quality of Life, summary components from the SF-36;
SE: standard error.

**Table 2b:**
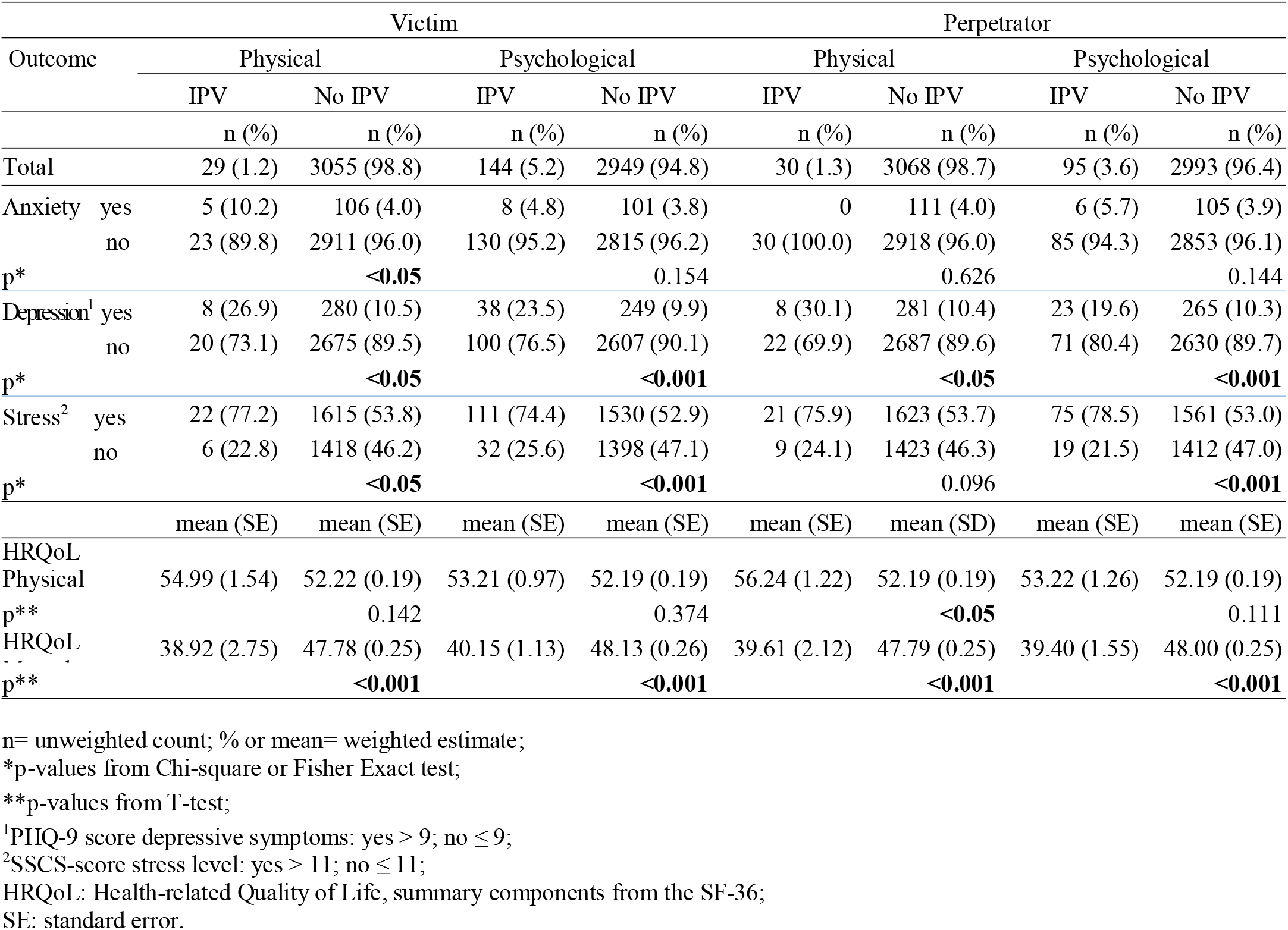
Mental health outcomes of women involved in intimate partner violence (IPV) as victims and perpetrators compared to women not involved in IPV according to type of violence (physical, psychological)

**Table 3:** Associations of physical and psychological intimate partner violence (IPV) and anxiety disorders, current depressive symptoms (PHQ-9)^1^ and stress (SSCS)^2^, in victims and perpetrators compared to participants not involved in IPV, stratified by sex

|  |  | Anxiety |  | Depression<sup>1</sup> |  | Stress<sup>2</sup> |  |
| --- | --- | --- | --- | --- | --- | --- | --- |
|  |  | Men | Women | Men | Women | Men | Women |
|  |  | aOR*<br>(95%-CI) | aOR*<br>(95%-CI) | aOR*<br>(95%-CI) | aOR*<br>(95%-CI) | aOR*<br>(95%-CI) | aOR*<br>(95%-CI) |
| Involvement |  |  |  |  |  |  |  |
| Physical IPV Victim | no | 1 | 1 | 1 | 1 | 1 | 1 |
|  | yes | 12.23<br>(2.29-65.40) | 3.74<br>(0.94-14.79) | 3.72<br>(0.65-21.42) | 2.31<br>(0.75-7.14) | 6.21<br>(1.42-27.28) | 2.66<br>(0.89-7.93) |
| Psychological IPV Victim | no | 1 | 1 | 1 | 1 | 1 | 1 |
|  | yes | 1.40<br>(0.40-4.84) | 1.15<br>(0.37- 3.56) | 3.85<br>(1.87-7.92) | 1.77<br>(0.92-3.39) | 3.92<br>(1.98-7.73) | 2.04<br>(1.23-3.38) |
| Physical IPV Perpetrator | no | - | - | 1 | 1 | 1 | 1 |
|  | yes | - | - | 1.98<br>(0.35-11.35) | 3.72<br>(1.34-10.30) | 4.74<br>(0.79-28.45) | 2.46<br>(1.04-5.86) |
| Psychological IPV Perpetrator | no | 1 | 1 | 1 | 1 | 1 | 1 |
|  | yes | 3.08<br>(0.89-10.68) | 1.58<br>(0.46-5.39) | 1.27<br>(0.49-3.30) | 2.14<br>(0.90-5.06) | 5.49<br>(2.57-11.73) | 2.76<br>(1.33-5.70) |

<sup>1</sup>depressive symptoms according to PHQ-9 scores: yes > 9; no ≤ 9;
<sup>2</sup>stress level according to SSCS score: “below-average to average” ≤ 11; “above-average” ≥ 12;
\*models adjusted for age group, marital status, SES, general health status and childhood abuse; OR: Odds Ratio; 95%CI: 95% Confidence Intervals;
1: reference group;
Models fitted using Complex Samples Logistic Regression.

**Table 4:** Associations between physical and psychological intimate partner violence (IPV) and the physical and mental summary components of health-related quality of life (SF-36) in victims and perpetrators, stratified by sex

| QoL | Involvement |  | Men | Women |
| --- | --- | --- | --- | --- |
| | | | $\beta$ ** (SE) | $\beta$ ** (SE) |
| Physical QoL | Physical IPV Victim | no | 1 | 1 |
|  |  | yes | -1.789 (2.281) | 1.273 (1.269) |
|  |  | [95% CI] | [-6.289; 2.711] | [-1.231; 3.777] |
| Mental QoL | Physical IPV Victim | no | 1 | 1 |
|  |  | yes | -4.667 (1.329)* | -6.905 (2.599)* |
|  |  | [95% CI] | [-7.290; -2.045] | [-12.033; -1.777] |
| Physical QoL | Psychological IPV Victim | no | 1 | 1 |
|  |  | yes | -1.164 (1.118) | 0.247 (0.966) |
|  |  | [95% CI] | [-3.370; 1.041] | [-1.660; 2.154] |
| Mental QoL | Psychological IPV Victim | no | 1 | 1 |
|  |  | yes | -6.772 (1.270)* | -6.426 (1.181)* |
|  |  | [95% CI] | [-9.279; -4.266] | [-8.757; -4.095] |
| Physical QoL | Physical IPV Perpetrator | no | 1 | 1 |
|  |  | yes | 1.705 (2.605) | 2.434 (1.181)* |
|  |  | [95% CI] | [-3.436; 6.846] | [0.103; 4.764] |
| Mental QoL | Physical IPV Perpetrator | no | 1 | 1 |
|  |  | yes | -5.625 (3.122) | -7.231 (2.292)* |
|  |  | [95% CI] | [-11.787; 0.536] | [-11.753; -2.708] |
| Physical QoL | Psychological IPV Perpetrator | no | 1 | 1 |
|  |  | yes | 2.536 (3.046)* | 0.449 (1.146) |
|  |  | [95% CI] | [0.924; 4.148] | [-1.813; 2.711] |
| Mental QoL | Psychological IPV Perpetrator | no | 1 | 1 |
|  |  | yes | -5.379 (1.431)* | -7.769 (1.623)* |
|  |  | [95% CI] | [-8.202; -2.556] | [-10.971; -4.567] |

\*p-value < 0.05;
\*\*adjusted for age group, marital status, SES and childhood abuse;
1: reference group;
SE: standard error;
Models fitted using Complex Sample General Linear Model.

